# The detrimental effect of stroke on motor adaptation

**DOI:** 10.1101/2024.03.21.24304376

**Authors:** Sabrina J. Abram, Jonathan S. Tsay, Heran Yosef, Darcy S. Reisman, Hyosub E. Kim

**Affiliations:** Department of Psychology, University of California, Berkeley; Helen Wills Neuroscience Institute, University of California, Berkeley; Department of Psychology, Carnegie Mellon University; Department of Physical Therapy, University of Delaware; School of Kinesiology, University of British Columbia; Graduate Program in Neuroscience, University of British Columbia

**Keywords:** stroke, motor control, motor adaptation, motor impairment, systematic review, meta-analysis

## Abstract

**Background:** While it is evident that stroke impairs motor control, it remains unclear whether stroke impacts motor adaptation—the ability to flexibly modify movements in response to changes in the body and the environment. The mixed results in the literature may be due to differences in participants’ brain lesions, sensorimotor tasks, or a combination of both.

**Objective:** We first sought to better understand the overall impact of stroke on motor adaptation and then delineated the impact of lesion hemisphere and sensorimotor task on adaptation poststroke.

**Methods:** Following the Preferred Reporting Items for Systematic reviews and Meta-Analyses (PRISMA) guidelines, we conducted a systematic review and meta-analysis of 17 studies comparing individuals poststroke to neurotypical controls, with each group consisting of over 200 participants.

**Results:** We found that stroke impairs motor adaptation (d = −0.64; 95% CI [−1.06, −0.22]), and that the extent of this impairment did not differ across sensorimotor tasks but may vary with the lesioned hemisphere. Specifically, we observed greater evidence for impaired adaptation in individuals with left hemisphere lesions compared to those with right hemisphere lesions.

**Conclusions:** This review not only clarifies the detrimental effect of stroke on motor adaptation but also underscores the need for finer-grained studies to determine precisely how various sensorimotor learning mechanisms are impacted. The current findings may guide future mechanistic and applied research at the intersection of motor learning and neurorehabilitation.

## Introduction

It is indisputable that stroke impairs motor control, as evidenced by symptoms like hemiparesis, rigidity, and loss of independent joint control.^1–10^ However, it is still unclear whether stroke impairs motor adaptation, the ability to flexibly respond to changes in the body (e.g., muscle fatigue) and the environment (e.g., walking on uneven terrain). The results of studies examining motor adaptation in individuals poststroke are mixed, with some finding impaired adaptation,^11^ while others have found no impairments.^12^

These mixed results may be due to the heterogeneity of lesion locations, experimental tasks, or a combination of both. Given that the right and left hemispheres appear to contribute differently to cognition^13^ and motor control,^14^ they may also be differentially involved in adaptation. Indeed, there is evidence pointing towards the selective involvement of the left hemisphere in adaptation.^15^ And given that different experimental tasks may rely on different mechanisms,^16^ the impact of a stroke may also differ between tasks that involve the upper limb (e.g., visuomotor adaptation) and those that involve the lower limb (e.g., split-belt adaptation). The time is ripe to comprehensively evaluate how the lesioned hemisphere and experimental task affect motor adaptation outcomes poststroke.

Here, we performed a meta-analysis to gain a better understanding of the effect of stroke on motor adaptation. By synthesizing adaptation behavior across 17 studies involving over 200 individuals poststroke, we determined the degree to which stroke impairs adaptation. By conducting subgroup analyses, we determined whether the level of impairment was influenced by lesion hemisphere and experimental task. Together, these findings may guide future mechanistic and applied research at the intersection of motor learning and neurorehabilitation.

## Methods

### Study selection criteria

We defined four criteria for determining whether studies were included in this meta-analysis: (1) studies included data from individuals poststroke as well as age-matched neurotypical controls; (2) outcome measures included those associated with motor adaptation; (3) measures of adaptation were collected during the perturbation block, and preferably, immediately after the perturbation was removed; and (4) studies were written in English. At every stage of this systematic review, we adhered to the Preferred Reporting Items for Systematic reviews and Meta-Analyses (PRISMA) guidelines.^17^

### Article screening

We identified and screened articles from several large databases including the Association for Computing Machinery Digital Library (ACM), the Cumulative Index to Nursing and Allied Health Literature (CINAHL), the Cochrane Central Register of Controlled Trials (CENTRAL), ProQuest, PubMed, and Scopus (Figure 1). We used the following search terms: implicit OR explicit OR upper extremity OR lower extremity OR paretic OR non-paretic OR dominant OR non-dominant OR subcortical OR cortical OR cortex AND motor learning AND stroke. For studies that involved the lower extremity, we narrowed our inclusion criteria to focus specifically on the most common task: split-belt walking. We also solicited articles from social media, tables of contents from relevant journals (e.g., NNR), and citations from related systematic reviews.^18,19^

**Figure 1.**
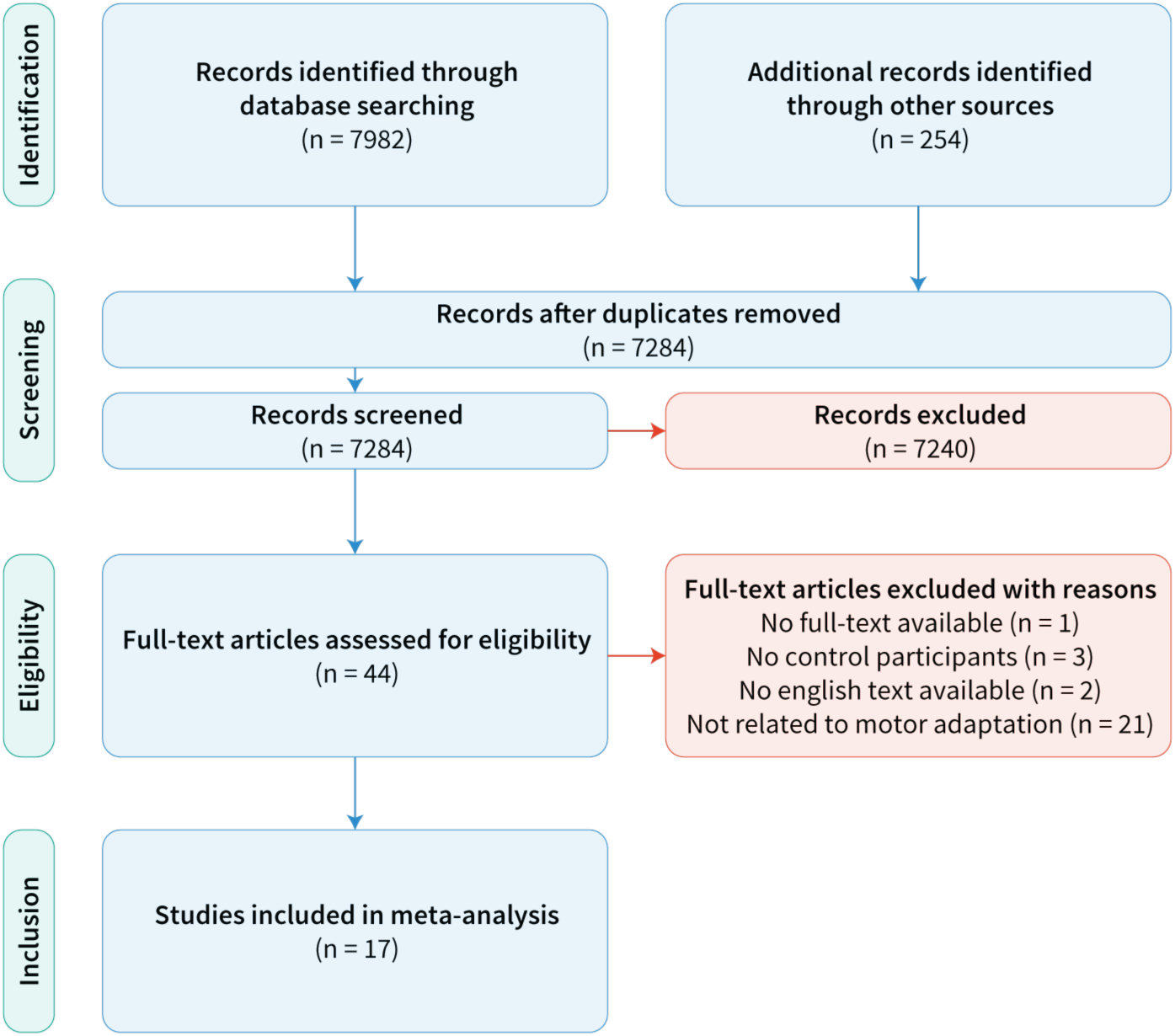
PRISMA flow diagram describing the study inclusion process of the systematic review. We identified 17 studies, resulting in 20 total datasets, that fulfilled our eligibility criteria.

We included 20 datasets from 17 studies in our meta-analysis (Figure 1). We identified 8236 studies using the search terms in the above databases and sources. Two authors removed 952 articles that were duplicates and an additional 7240 articles based on the title and abstract. All authors then independently inspected the full text of 44 remaining articles based on our four eligibility criteria and agreed upon which articles should be included. We removed one study that did not have full text available, three that did not include age-matched neurotypical participants, two that were not written in English, and 21 that were outside the scope of motor adaptation. Of the 17 remaining studies, 15 were published in peer-reviewed journals and two were unpublished datasets. All data used for this meta-analysis are available on OSF (https://osf.io/gfmdr/).

From the 20 eligible datasets, we extracted the following: 1) sample size and average age of patients and controls; 2) lesion hemisphere and lesion location; 3) whether the intact limb or paretic limb was used in the behavioral task; and 4) motor adaptation, that is, the adaptive changes in behavior either during the perturbation block or the aftereffect block immediately after the perturbation was removed. We outline below how we standardized motor adaptation outcomes. In total, there were 247 participants with stroke and 208 controls. The sample sizes for independent datasets were relatively small (stroke: n = 2-27; control: n = 5-31), further motivating a meta-analysis approach to synthesize data across the literature.

### Data synthesis and analysis

We analyzed a range of motor adaptation tasks including those that involved saccades, reaching, and walking. In reaching tasks, there were three types of paradigms that varied in the nature of the perturbation: in visuomotor rotation tasks, the participants reached to a visual target and received feedback in the form of a visual cursor whose radial distance was matched to the hand but angular distance rotated with respect to the hand (green; Figure 2A);^20–25^ in visuomotor gain tasks, the visual cursor was perturbed along the radial dimension, while the angular distance was matched to that of the hand (red; Figure 2A);^26^ and in force field adaptation tasks, the participants reached to a target with a robot arm applying forces to the hand (orange; Figure 2A).^27^ In saccade adaptation tasks, the participants made goal-directed eye movements to targets, where the position of the target jumped immediately upon saccade initiation (yellow; Figure 2B).^28,29^ In gait adaptation tasks, the participants walked on a split-belt treadmill with the left and right legs moving at different speeds (purple; Figure 2C).^11,12,30–34^

**Figure 2.**
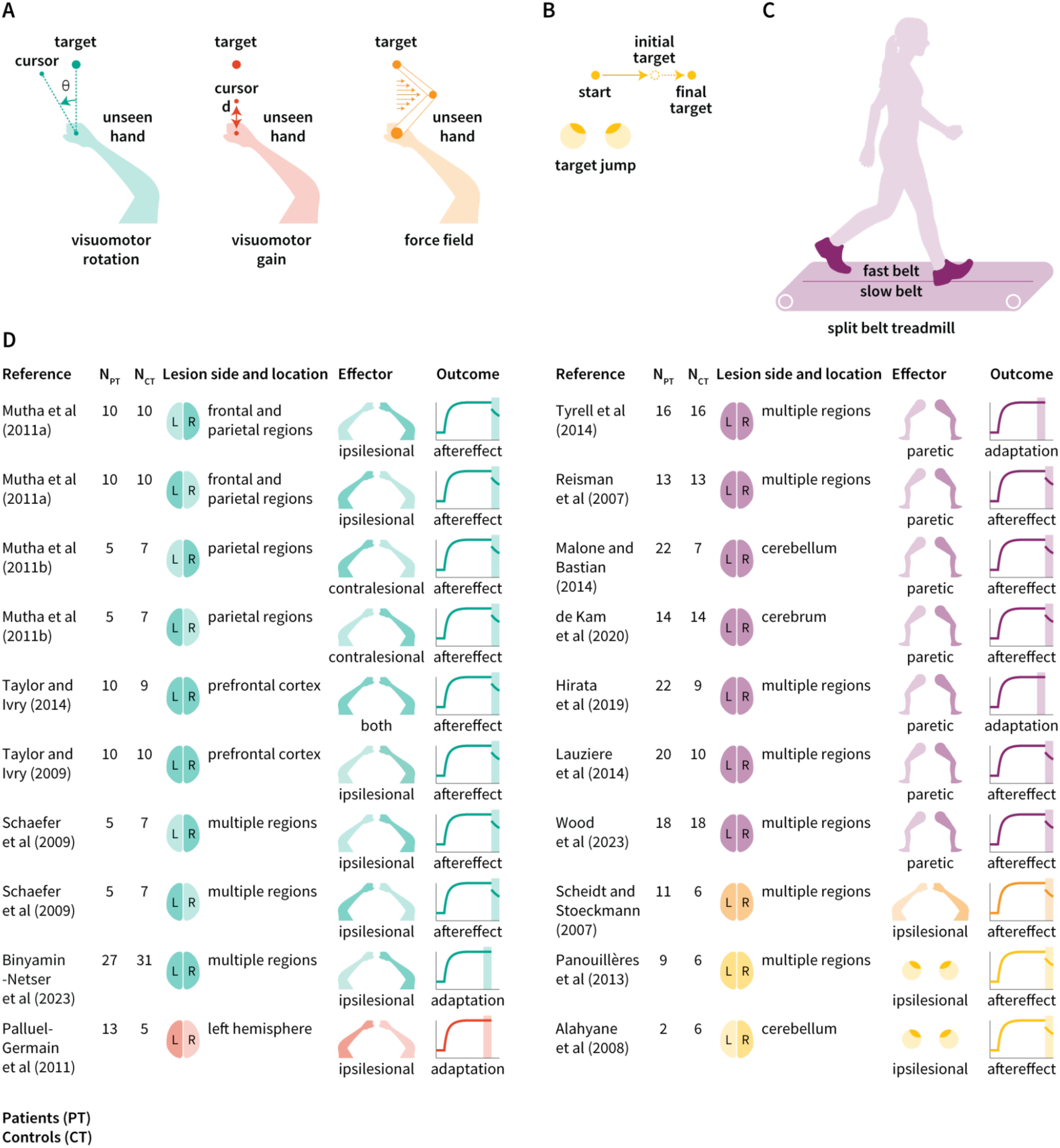
Overview of datasets included in this meta-analysis. Schematic of motor adaptation tasks involving (**A)** reaching, (**B)** saccades, and (**C)** walking. (**D)** The number of patients (PT) and controls (CT), lesion side and location, limb used by participants poststroke (effector), and outcome measure for each dataset. Darker shading indicates lesion hemisphere (left, right, or both, meaning that the dataset includes individuals with both left and right hemisphere lesions), limb used (contralesional, ipsilesional, or both), and outcome measure (late adaptation or aftereffect).

Despite these differences in movements and perturbations, these tasks can be used to study motor adaptation. Mechanistically, the introduction of a perturbation causes sensory prediction errors, that is, a mismatch between predicted and actual sensory outcomes of a movement. In visuomotor rotation tasks, for example, the sensory prediction errors are elicited by a discrepancy between the predicted cursor position and the actual cursor position. With experience, people learn to iteratively reduce this error by reaching away from the target in the direction opposite to the perturbation. When the perturbation is removed, there are often residual aftereffects in the same direction as learning. Given that motor adaptation refers to changes in feedforward control (i.e., how future movements are planned and executed), aftereffects are often regarded as the key signature of learning since it is not influenced by online feedback corrections made during the movement itself.

We focused our meta-analysis on two possible time points: late adaptation (i.e., towards the end of the perturbation block) or aftereffects (i.e., immediately after the removal of the perturbation). Of the 20 datasets included in this meta-analysis, 4 measured adaptation only during the perturbation block whereas 16 measured both adaptation during the perturbation block and aftereffects (Figure 2D). We standardized measures across datasets with different dependent variables (e.g., hand angle, step length asymmetry, and saccade amplitude) by using the mean and standard deviation for patients and controls to calculate Cohen’s d and 95% confidence intervals (CI). If the mean and standard deviation were not reported, we either used F-or t-statistics that compared performance between groups,^35^ or estimated these values using the GRABIT software (Doke, MATLAB Central File Exchange). If a study grouped participants into different categories (e.g., mild, moderate, and severe impairment), we estimated the combined mean and standard deviation. For example, in the case of two groups, we used the following equations:

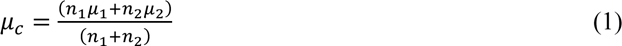

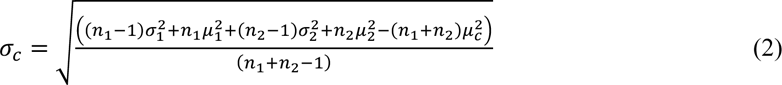

Where 𝑛 is the sample size, 𝜇 is the mean, 𝜎 is the standard deviation, a subscript of *c* represents the combined value, and subscripts of *1* or *2* represent the values for individual groups. We validated this method against simulated data.

We calculated the overall effect size using a random effects model, where the contribution of each dataset was weighted by the sample size and uncertainty in the effect size. We interpreted effect sizes less than 0.2 as small, between 0.2 and 0.8 as medium, and greater than 0.8 as large, and defined the significance level as 𝛼 = 0.05.^36^ To assess the heterogeneity or variability in effect sizes between datasets, we calculated the Q value, which compares the weighted effect sizes to the overall effect size. Smaller Q values indicate less variance, while larger Q values indicate more variance. To determine whether this variability is greater than that expected by chance, we used Cochran’s Q test, which compares the calculated Q value to a chi-squared distribution with degrees of freedom equal to the number of datasets minus one. A significant result (p < 0.05) indicates heterogeneity among effect sizes, meaning the datasets are not consistent with each other. All statistical analyses were performed using R and RStudio (code available at https://osf.io/gfmdr/).

### Subgroup analyses

We conducted subgroup analyses related to two covariates: lesion hemisphere and experimental task (Figure 2D). To determine the effect of lesion hemisphere, we compared the five datasets that tested individuals with unilateral stroke in the left hemisphere (n = 43) to the five datasets that tested individuals with unilateral stroke in the right hemisphere (n = 30). This analysis did not include datasets that combined individuals with left and right hemisphere stroke (n = 174). Note that the Wood et al. 2023 study did not intend to compare laterality differences. However, given that each participant with stroke was yoked to an age-matched control and we had this information as well as information regarding the affected hemisphere for each participant, we chose to include these data in this analysis. Next, to determine the effect of experimental tasks, we compared nine visuomotor rotation datasets (n = 87) with seven split-belt walking datasets (n = 125), as these two tasks made up the majority of datasets. We also conducted subgroup analyses only in cases where there were four or more datasets. When testing for differences between subgroups, we applied the same approach as when testing for differences between individual datasets. That is, we calculated the Q value comparing the subgroup effect sizes to the overall effect size. To determine whether variability between subgroups (left vs. right hemisphere lesions or visuomotor rotation vs. splitbelt walking tasks) is greater than chance, we used Cochran’s Q test with degrees of freedom equal to the number of subgroups minus one.

## Results

### The detrimental effect of stroke on motor adaptation

Across 20 datasets, motor adaptation was impaired in individuals poststroke compared to age-matched neurotypical controls (d = −0.64; 95% CI [−1.06, −0.22]; t_19_ = −3.20; p = 0.0047; Figure 3A and 3B). To test the reliability of this finding, we repeated this analysis after excluding one outlier (Lauziere et al. 2014) and again found that motor adaptation was impaired in individuals poststroke (d = −0.49; 95% CI [−0.74, −0.25]; t_18_ = −4.25; p = 0.00048). We next repeated this analysis for late adaptation, taking into consideration that, in some paradigms (i.e., split-belt adaptation), the aftereffect measurement may depend on the initial perturbation, the magnitude of which is not always the same in stroke patients and controls. We found that late adaptation was also impaired in individuals poststroke (d = −0.44; 95% CI [−0.85, −0.029]; t_19_ = −2.24; p = 0.037; Figure S1A and S1B). Lastly, we found that effect sizes varied across these 20 datasets with I^2^ = 62.4% (heterogeneity based on 𝜒^2^: Q_19_ = 50.54; p = 1.09 x 10^-4^), further motivating our subgroup analyses.

**Figure 3.**
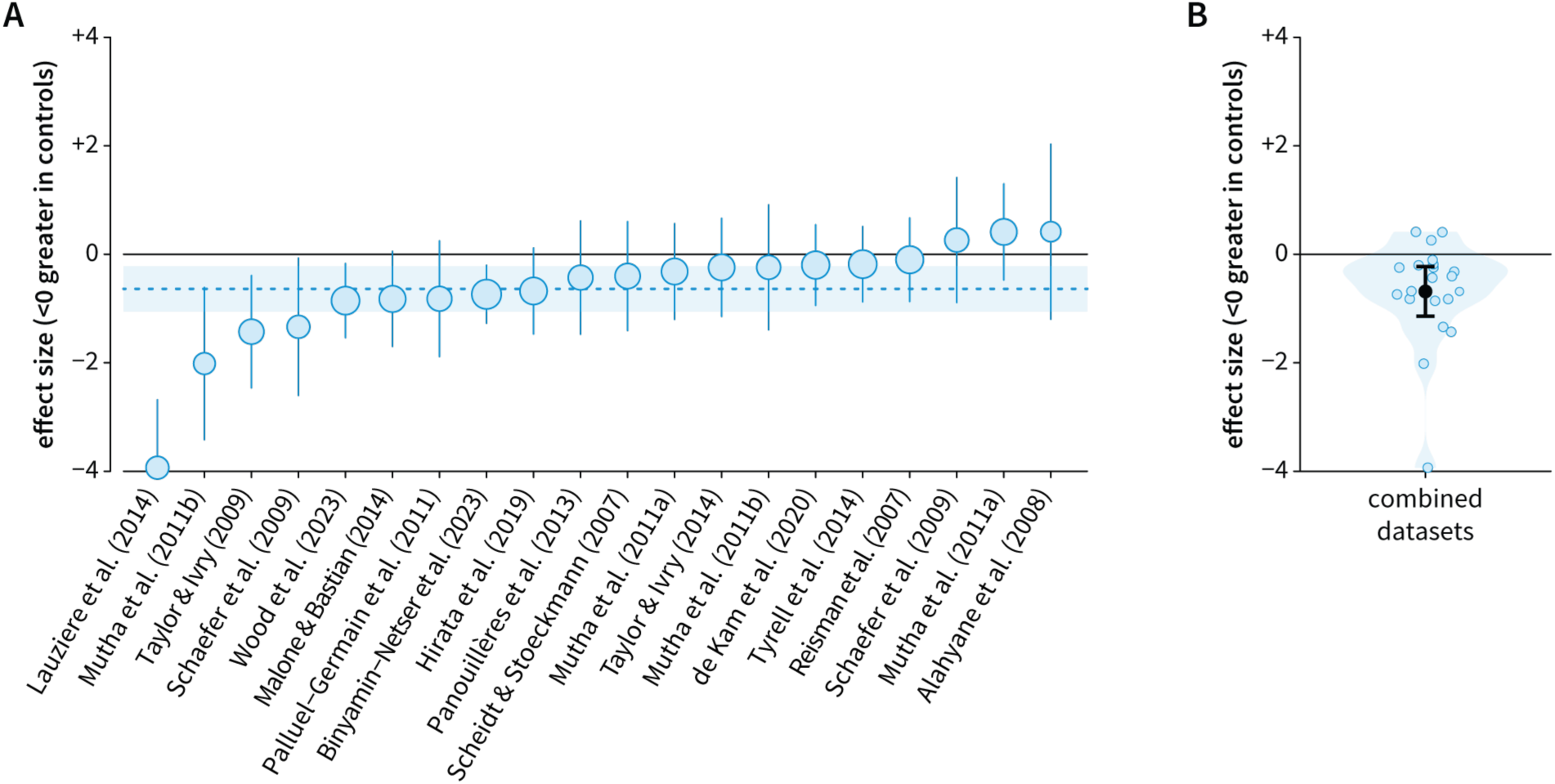
Stroke impairs motor adaptation. **A.** Forest plot comparing the performance of individuals poststroke to neurotypical controls, where negative values indicate greater adaptation in controls (i.e., impaired adaptation poststroke). The overall effect size is indicated by the blue dashed line, and the 95% confidence interval is indicated by the blue shading. Each circle represents a single dataset with its size indicating the weight assigned to that dataset in the random-effects model. Whiskers represent the 95% confidence intervals. **B.** Violin plot displaying the distribution of effect sizes. Each blue dot represents a single dataset, the black dot represents the mean across datasets, and the black whiskers represent the standard error.

### The impact of experimental tasks on measures of motor adaptation poststroke

We did not find any significant differences in motor adaptation poststroke between visuomotor rotation and split-belt walking tasks (Q_1_ = 0.39; p = 0.53), suggesting that differences between tasks are not the underlying driver of mixed results in the literature. When examining each task in isolation, the results were less robust as compared to our analysis of all tasks combined. However, we still observed medium-to-large effect sizes related to the negative impact of stroke on adaptation in both visuomotor rotation (d = −0.56; 95% CI [−1.14, +0.019]; t_8_ = −2.23; p = 0.056; Figure 4A and 4B) and split-belt walking (d = −0.90; 95% CI [−2.08, +0.28]; t_6_ = −1.87; p = 0.11; Figure 4A and 4B) tasks.

**Figure 4.**
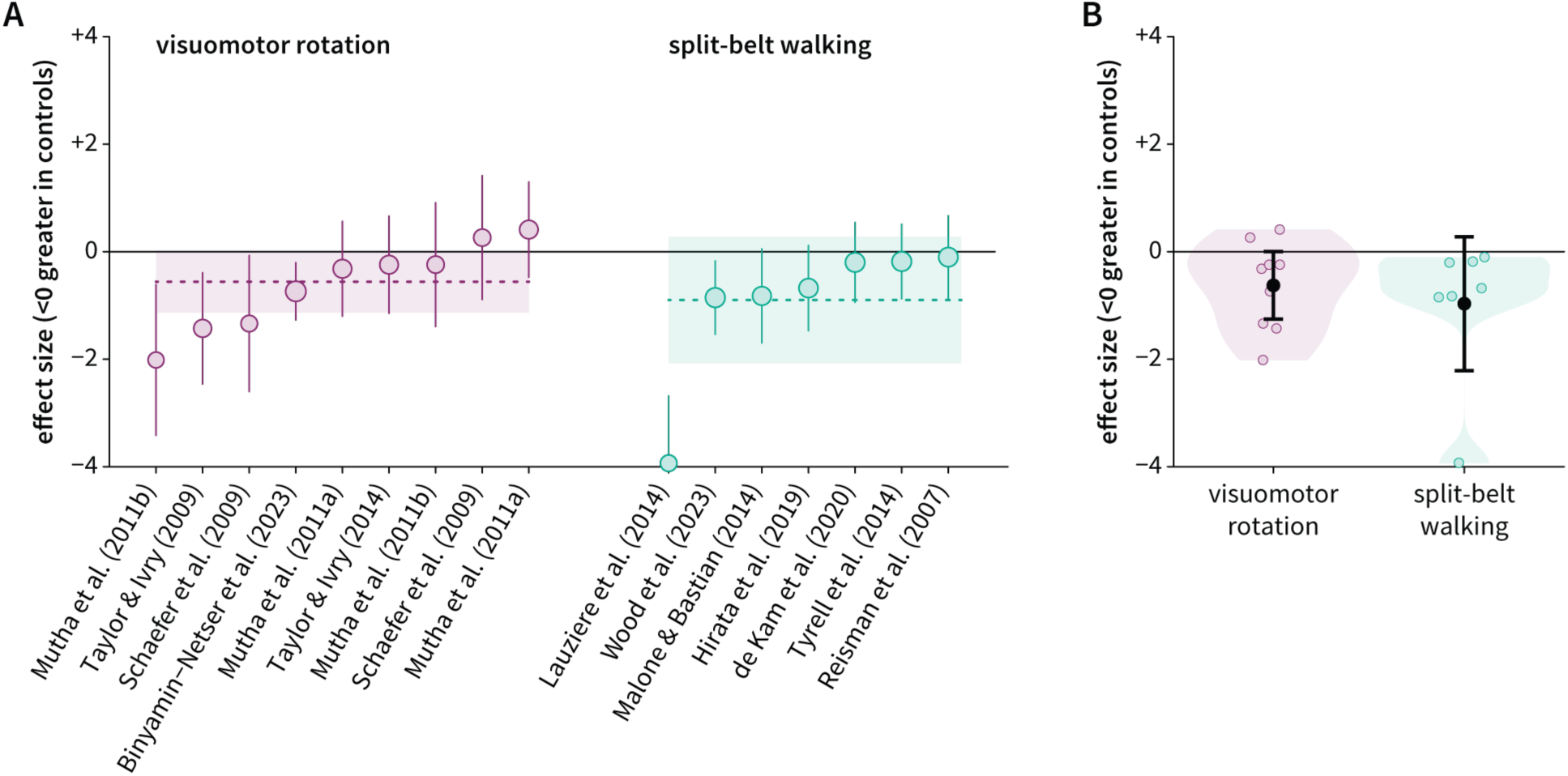
Minimal impact of experimental tasks on motor adaptation poststroke. **A.** We assigned subgroups based on whether datasets used visuomotor rotation or split-belt walking tasks and then compared performance of individuals poststroke to controls in each subgroup. The effect size for each subgroup (purple for visuomotor rotation and green for split-belt walking) is indicated by the dashed line and 95% confidence intervals by the shading. Each circle represents a single dataset with its size indicating its weight and whiskers representing 95% confidence intervals. **B.** Violin plot comparing the distribution of effect sizes between datasets using visuomotor rotation (purple) and split-belt walking (green). The black dot represents the mean value for each subgroup and the black whiskers represent the standard error.

### The impact of lesion hemisphere on measures of motor adaptation poststroke

While nearly all studies in this meta-analysis tested individuals with unilateral stroke, many did not examine the impact of lesion hemisphere on motor adaptation (i.e., left vs. right hemisphere lesions). When we examined datasets that did report the lesioned hemisphere and provide corresponding results, we found that motor adaptation was selectively impaired in individuals with left hemisphere lesions (d = −0.79; 95% CI [−1.61, +0.027], t_4_ = −2.69, p = 0.055; Figure 5A and 5B) but intact in individuals with right hemisphere lesions (d = −0.14; 95% CI [−1.14, +0.86], t_4_ = −0.39, p = 0.71; Figure 5A and 5B). When we performed a direct comparison of motor adaptation with left vs. right hemisphere lesions, we observed an effect of hemisphere (d = −1.11; 95% CI [−2.68, 0.45]), albeit one that was not robust (Q_1_ = 1.97, p = 0.16). One reason for this lack of robustness may be low statistical power with only five datasets in each group. Another reason may be that other covariates, such as brain regions within each hemisphere and the limb used during the task, influenced adaptation measures (see Discussion). Despite these caveats, these results hint toward the left hemisphere being a neural locus for motor adaptation.

**Figure 5.**
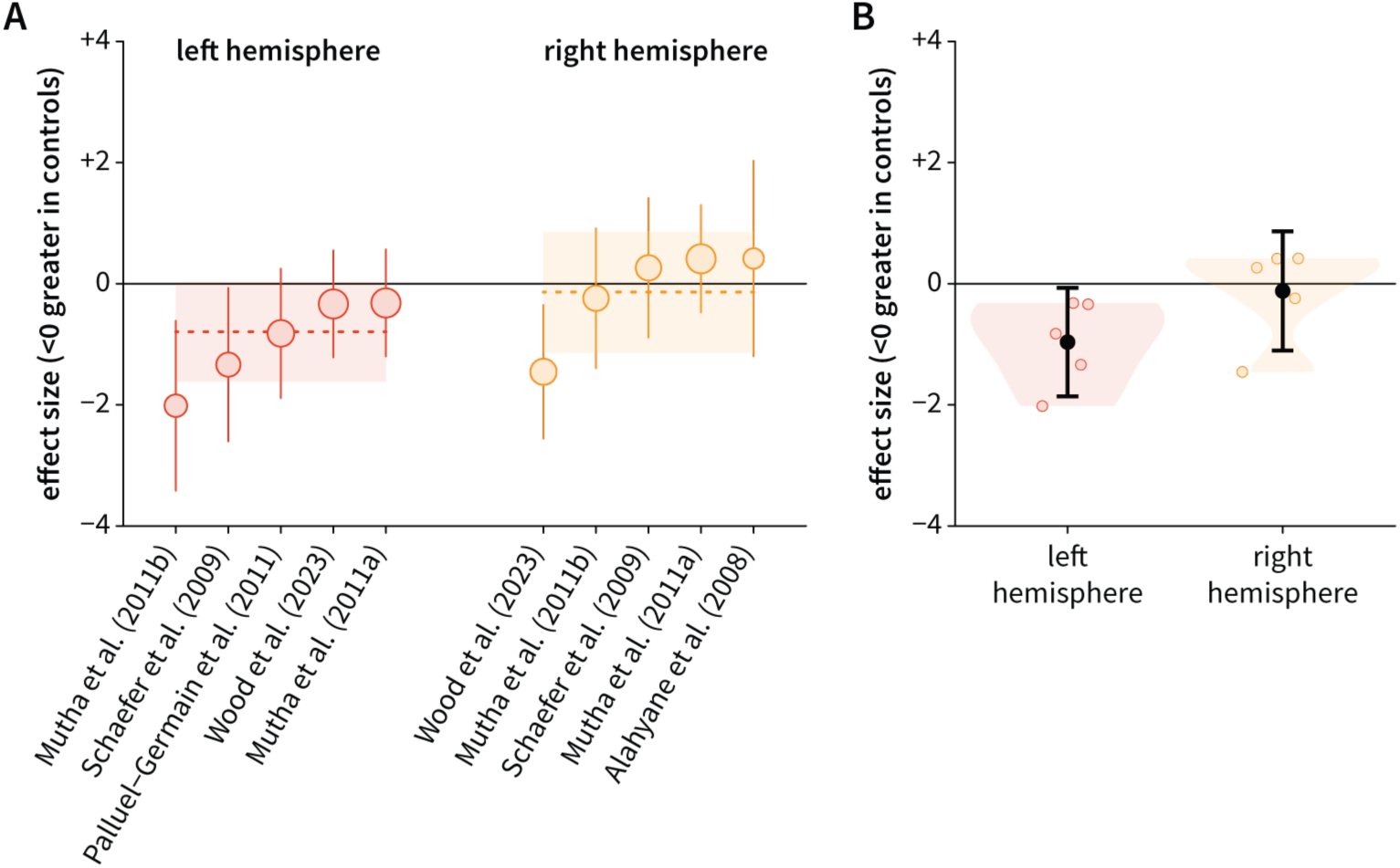
Motor adaptation is impaired in individuals with left hemisphere lesions but intact in individuals with right hemisphere lesions. **A.** Subgroups were assigned based on whether datasets compared motor adaptation between individuals with left hemisphere lesions or right hemisphere lesions against controls. The effect size for each subgroup (red for left hemisphere and orange for right hemisphere) is indicated by the dashed line and 95% confidence intervals by the shading. Each circle represents a single dataset with its size indicating its weight and whiskers representing 95% confidence intervals. **B.** Violin plot comparing the distribution of effect sizes between datasets examining left hemisphere lesions (red) and right hemisphere lesions (orange). The black dot represents the mean value for each subgroup and the black whiskers represent the standard error.

## Discussion

We conducted a comprehensive review to examine the impact of stroke on motor adaptation. By synthesizing 20 datasets involving over 200 individuals poststroke, we found that motor adaptation was impaired in individuals poststroke compared to neurotypical controls. Notably, impairments in motor adaptation were consistent across various sensorimotor tasks. In contrast to this consistency across paradigms, our initial results reveal that these impairments may be influenced by lesioned hemisphere: motor adaptation was impaired in individuals with left hemisphere lesions but intact in individuals with right hemisphere lesions. This meta-analysis not only highlights the detrimental effect of stroke on motor adaptation but also hints toward the left hemisphere serving as a critical locus for this process.

### Left hemisphere as a neural correlate of motor adaptation

Our meta-analysis points toward the left hemisphere as a neural correlate of motor adaptation. This finding aligns with the notion that the left hemisphere primarily contributes to feedforward control, while the right hemisphere contributes more to feedback control.^14,15^ Another possibility is that the left hemisphere is critical for logical reasoning, which may be needed for the development of a successful explicit strategy, particularly in visuomotor adaptation tasks (refer to next section for methods that effectively isolate implicit recalibration and explicit strategies).^37,38^ Given limited studies that control for lesion hemisphere (and lesion location), future studies using methods such as lesion-symptom mapping are needed to precisely determine the contribution of these factors to motor adaptation.^39^

### Investigating the impact of stroke on implicit recalibration and explicit strategies

Our meta-analysis not only clarified the detrimental impact of stroke on motor adaptation but also identified an important gap in our understanding: *how* does stroke impact the different learning processes supporting motor adaptation? Here, we focus on upper extremity tasks like visuomotor adaptation as it is now well established that multiple learning processes support changes in this behavior.^40–43^ There is more work to be done to determine whether these same processes contribute to adaptation in lower extremity tasks such as split-belt walking. For these upper extremity tasks, two of the most salient processes include *implicit recalibration*, which keeps our motor system well-calibrated in a gradual and subconscious manner, and *explicit strategies*, which, in contrast, enables rapid and volitional motor corrections.^38^ Existing studies have not cleanly examined the impact of stroke on each process.

There are many experimental tools for dissociating the processes that underlie adaptation.^38,44–49^ These tools will be especially useful for identifying how stroke impacts these processes. The first method for isolating implicit recalibration involves clearly instructing participants immediately prior to aftereffect trials, in which the perturbation is removed, to forgo any explicit strategies they may have been using (Figure 6A). While most of the studies included in this meta-analysis measured aftereffects, we could not ascertain whether proper instructions were provided (e.g.., “Move your hand directly to the target and do not aim away from the target.”). Interestingly, a recent study that provided proper instructions found no effect of stroke on implicit recalibration.^25^ Given the singular nature of this study, it will be enlightening to re-examine how stroke impacts this implicit process after several more studies adopt this approach. The second approach is to use clamped visual feedback.^38,46^ Unlike standard visuomotor rotation tasks, the clamped visual cursor moves at a fixed angle away from the target and is not contingent on the participant’s hand angle (Figure 6A). Critically, we inform participants of this manipulation and instruct them to always reach directly to the target and ignore the clamped visual feedback. Despite these instructions, participants exhibit robust implicit recalibration, with learning occurring outside of conscious awareness.^50^

**Figure 6.**
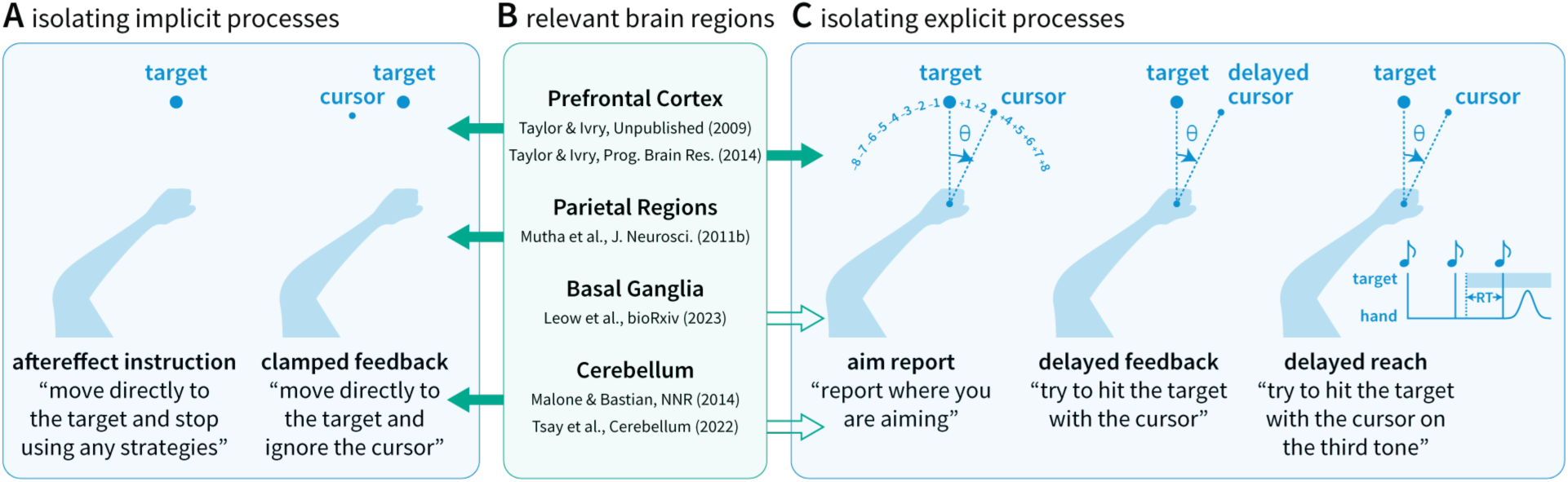
**A.** Methods for isolating implicit recalibration. **B.** Evidence suggesting which brain regions are involved in which learning processes.^20,22,24,31,65,66^ The solid arrows indicate studies that involve populations with stroke and open arrows indicate studies that involve populations with progressive neurodegenerative disorders. **C.** Methods for isolating explicit re-aiming strategies.

Paralleling the methodological advances for studying implicit recalibration, new approaches have been developed to examine explicit re-aiming strategies. One approach involves asking participants to indicate where they are aiming prior to each reach, revealing the dynamics of explicit re-aiming (Figure 6C).^22,50^ Another approach involves *delayed* rotated feedback, a manipulation that robustly attenuates implicit recalibration and thus isolates explicit re-aiming (Figure 6C).^51^ A final approach involves manipulations of preparation time. When manipulating the amount of time a visual target is presented before a reach begins, longer preparation time enables the use of explicit strategies that are resource-demanding, whereas shorter preparation time limits deliberation and minimizes the contribution of these explicit strategies (Figure 6C).^52–54^ Taken together, upper extremity tasks provide well-controlled methods for isolating implicit and explicit processes, which are either not present or not easily isolated in other experimental paradigms. Future studies using fine-grained experimental and computational methods to evaluate these processes in individuals poststroke will shed light on the neural correlates of different sensorimotor learning mechanisms (Figure 6B).

### From fundamental learning mechanisms to targeted rehabilitation strategies

A better understanding of how stroke affects motor adaptation will motivate targeted rehabilitation interventions that are tailored to the specific motor deficits and affected brain regions. Given that stroke impairs feedforward processes, rehabilitation programs could instead capitalize on the intact feedback processes. For example, reinforcement learning, the process of refining actions through reward or punishment feedback, may be an effective alternative.^61^ In addition, given that individuals with right hemisphere lesions appear to have intact feedforward processes, rehabilitation therapists might consider interventions that capitalize on this.^30,55,56^ Individuals with left hemisphere lesions, who may have feedforward deficits, could also benefit from exploiting existing neural resources via task-specific intensive training,^55,57,58^ or explore compensatory learning processes via reinforcement feedback.^2,59,60^ Regardless of the exact approach, we anticipate that finer-grained neuropsychological work will deepen our understanding of how stroke affects motor adaptation and inform future rehabilitation strategies aimed at repairing or remodeling affected neural circuits.^62–64^

## Data Availability

All data produced are available online at: https://osf.io/gfmdr/

## Supplemental Material

**Figure S1.**
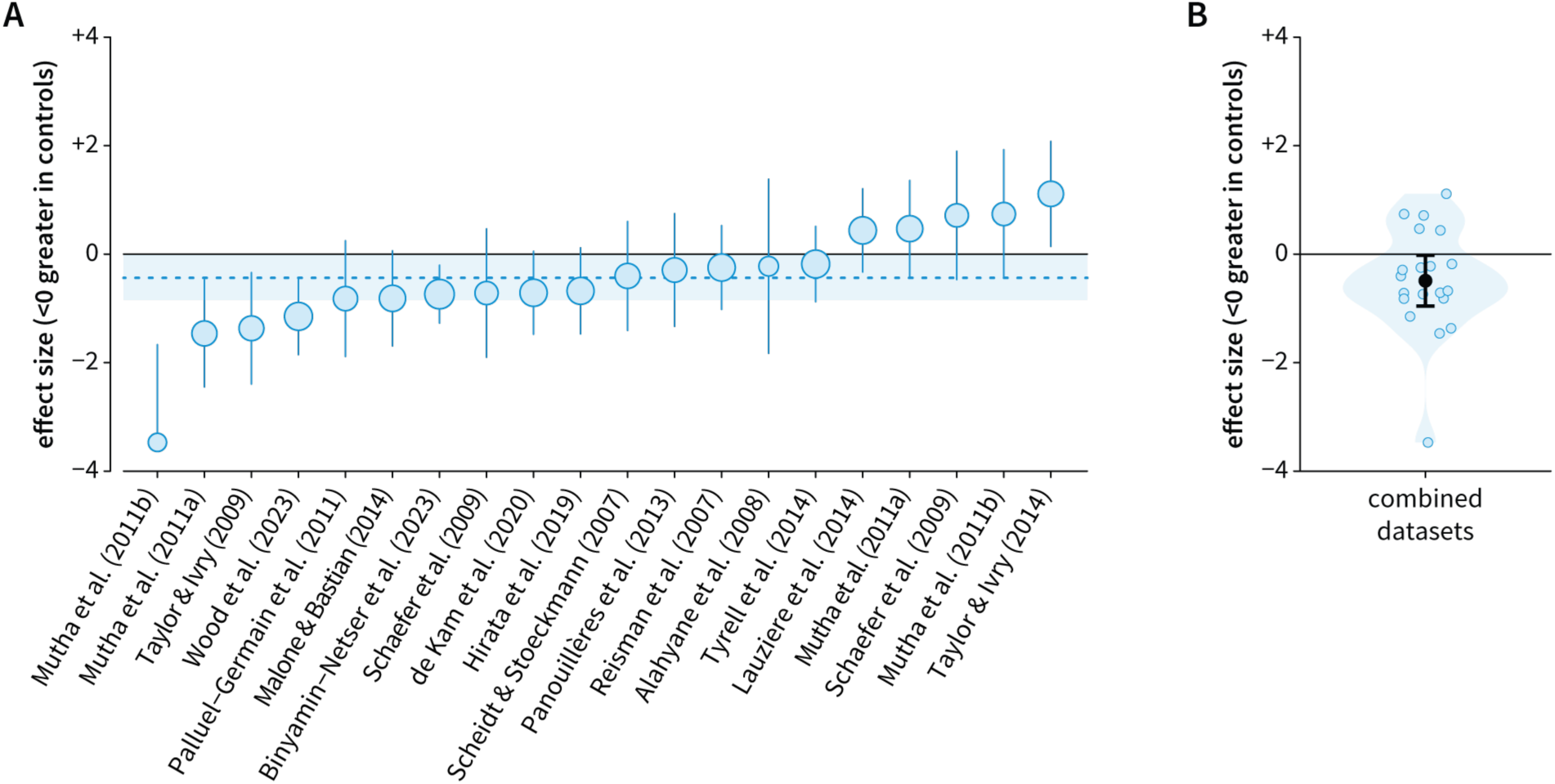
Late adaptation. **A.** Forest plot comparing the performance of individuals poststroke to neurotypical controls, where negative values indicate greater adaptation in controls (i.e., impaired adaptation poststroke). The overall effect size is indicated by the blue dashed line, and the 95% confidence interval is indicated by the blue shading. Each circle represents a single dataset with its size indicating the weight assigned to that dataset in the random-effects model. Whiskers represent the 95% confidence intervals. **B.** Violin plot displaying the distribution of effect sizes. Each blue dot represents a single dataset, the black dot represents the mean across datasets, and black whiskers represent the standard error.

## Notes

### Competing Interest Statement

The authors have declared no competing interest.

### Funding Statement

This study was funded by the National Institutes of Health

### Author Declarations

This work used only data that were openly available to the public before the initiation of the study.

